# Social and Leisure Activity Contribute to the Attenuation of Decline in Behavioral Variant Frontotemporal Degeneration

**DOI:** 10.1101/2021.01.10.21249399

**Authors:** Nikolas G. Kinney, Jessica Bove, Jeffrey S. Phillips, Katheryn A. Cousins, Christopher A. Olm, Daniel G. Wakeman, Corey T. McMillan, Lauren Massimo

**Affiliations:** Frontotemporal Degeneration Center, Perelman School of Medicine, Department of Neurology, Philadelphia, PA; School of Nursing, University of Pennsylvania, Philadelphia, PA

**Keywords:** Cognitive reserve, behavioral variant frontotemporal degeneration, social/leisure activity, lifetime of experiences questionnaire, cortical thickness

## Abstract

Behavioral variant frontotemporal degeneration (bvFTD) is clinically characterized by progressive decline in social and executive domains. Previous work suggests that early lifestyle factors such as education and occupational attainment may relate to structural integrity and moderate the rate of cognitive decline in bvFTD, but the role of other cognitively stimulating activities is understudied. We sought to investigate the effect of such activities on cortical thickness (CT) in bvFTD. bvFTD patients (n=31) completed a baseline MRI scan, and informants for the patients completed the Lifetime of Experiences Questionnaire (LEQ), which measures specific activities considered to be undertaken primarily within one particular life phase, such as education (young-life), occupation (mid-life), and social/leisure activity (late-life). At baseline, linear models assessed the effect of LEQ scores from each life phase on regional CT. A subset (n=19) of patients completed longitudinal MRI, and to evaluate the association of LEQ with longitudinal rates of CT decline, we derived individualized slopes of decline using linear mixed effects models and these were related to LEQ scores from each life phase. At baseline, a higher late-life LEQ score was associated with less atrophy in bilateral superior anterior temporal regions. Longitudinally, we observed that higher late-life LEQ scores were associated with an attenuated rate of CT loss in insular cortex. Late-life LEQ score was positively associated with both relatively preserved CT early in bvFTD and a slower rate of cortical loss in regions important for social functioning. These findings suggest that social and leisure activities may contribute to a form of resilience against pathologic effects of disease.

## 1. INTRODUCTION

Cognitive reserve (CR) has been proposed as a mechanism by which one sustains cognitive function in the face of neurodegenerative disease pathology, and this source of resilience has been proposed to account for differences in inter-individual variability in cognitive status (Stern, 2003). Specifically, CR posits that enriching lifetime experiences, such as education, occupational attainment, and social engagement, can provide protection against pathogenic factors of neurodegenerative disease (Stern, 2009). CR has been widely studied in Alzheimer’s Disease (AD), but less is known about how CR moderates the rate of longitudinal decline in other forms of dementia, such as bvFTD.

Behavioral variant frontotemporal degeneration (bvFTD) is a progressive neurodegenerative disease that primarily affects the frontal and temporal lobes and is associated with deficits in social and executive functioning (Rascovsky et al., 2011). The disease course of bvFTD is highly variable. Lifestyle factors have been proposed to contribute to individual differences in resilience to this progressive neurodegenerative disease, but the majority of investigation has focused on education and occupational attainment. Previous cross-sectional work indicates that higher levels of education and occupational attainment are related to frontal gray matter density and better performance in executive functioning (Placek et al., 2016) as well as longer survival (Massimo et al., 2015). Indeed, in a recent longitudinal investigation, decline in executive performance in individuals with higher occupational attainment was related to reduced gray matter density in frontal regions (Massimo et al., 2019), highlighting the importance of frontally-mediated mechanisms to reserve in bvFTD.

Although the majority of CR studies in bvFTD focus on education and occupational attainment, there is a need to study the potential reserve building effects of other lifetime activities, such as leisure time and social engagements, on progression of bvFTD. One of the primary impairments in bvFTD is social deficits, highlighting the critical need to study the effects of social engagement in the context of this disease. Social isolation has previously been associated with poor cognitive functioning in healthy aging (Evans et al., 2018), and lower social engagement is associated with an increased risk of AD (Hersi et al., 2017). Furthermore, leisure time activities have been shown to improve clinical outcomes in patients with genetic FTLD (Casaletto et al., 2020) – with more active lifestyles directly moderating the relationship between cortical atrophy and cognition. Therefore, we hypothesized that higher levels of social and leisure time activities would attenuate decline on MRI measures of gray matter in regions underlying social cognition in bvFTD. To test this hypothesis we examined the relationship between cortical thickness (CT) and measures of lifetime enrichment from the Lifetime of Experience Questionnaire (LEQ) (Valenzuela and Sachdev, 2007).

## 2. METHODS

### 2.1. Participants

Thirty-one patients who met inclusion/exclusion criteria (see below) and were clinically diagnosed with bvFTD using published criteria (Rascovsky et al., 2011) through weekly multidisciplinary consensus meetings by experienced clinicians (MG, DJI, LM) at the Penn Frontotemporal Degeneration Center (FTDC) were included in this study. Participant demographics and characteristics are described in Table 1. As previous work has shown that the effect of CR may differ in autopsy confirmed FTLD and AD (Massimo et al., 2015), we constrained our bvFTD cohort to cases with likely FTLD pathology; exclusion criteria included a cerebrospinal fluid total tau:amyloid-beta cutoff of ≥0.34 which has >95% accuracy for identifying Alzheimer’s pathology (Irwin et al., 2012). To focus our study on social/behavioral impairments, we additionally excluded patients with a concurrent clinical diagnosis of a motor syndrome such as amyotrophic lateral sclerosis, corticobasal syndrome, or progressive supranuclear palsy, due to their more heterogeneous distributions of cortical atrophy and clinical decline (Armstrong et al., 2013; Irwin et al., 2013; Litvan et al., 1996; Rohrer et al., 2011; Strong et al., 2009; Whitwell et al., 2010). Medical and psychiatric causes of dementia were excluded by clinical exam, blood, and brain imaging tests.

**Table 1.** Participant Baseline Demographics and Clinical Characteristics

| | bvFTD | Control | $\chi^2/W$ | P |
| --- | --- | --- | --- | --- |
| Sex N (% Female) | 31 (29%) | 31 (32%) | 0 | 1 |
| Age: Median (IQR) | 62 (59.0-67.0) | 66 (59.5-71.5) | 540.5 | 0.40 |
| Dx Duration: Median (IQR) | 3.58 (2.42-5.42) | N/A | N/A | N/A |
| Education: Median (IQR) | 18 (16-18) | 18 (16-18) | 495 | 0.84 |
| MMSE: Median (IQR) | 25.5 (21-28) | 29 (29-30) | 812 | <0.05 |

Legend: M = Male, F = Female, IQR = Interquartile Range

Inclusion criteria included completion of the LEQ and an MRI scan. A subset of patients (n=19) additionally participated in longitudinal MRI. Eleven participants had one follow-up scan available, and eight had two. Median time for follow-up MRI was 1.08 years (range=9-36 months). We previously reported clinical and MRI data for two of these patients in a study of occupational attainment in bvFTD (Massimo et al., 2019).

To identify areas of cortical atrophy in bvFTD patients we developed a matched cohort of healthy control participants. Inclusion criteria for healthy controls were a self-report of no history of neurologic or psychiatric disease and a Mini-Mental State Examination score ≥27 (Folstein et al., 1975). From this pool of controls (N=97) we then used a propensity model strategy with the R package MatchIt (Ho et al., 2007) to identify a ratio of one patient to one matched control based on age, sex, and education. This resulted in a demographically matched cohort with no significant difference in age, sex, or education as measured by either chi-squared test (for sex) or non-parametric Mann-Whitney-Wilcoxon tests (see Table 1).

All participants and/or legally responsible representatives participated in an informed consent procedure approved by an Institutional Review Board convened at the University of Pennsylvania.

### 2.2. Lifetime of Experience Questionnaire

The LEQ (Valenzuela and Sachdev, 2007) was completed by a knowledgeable caregiver informant (typically spouse) for each patient. The LEQ, an instrument that assesses cognitive lifestyle (Valenzuela and Sachdev, 2007), has been cited as a comprehensive CR proxy measure (Kartschmit et al., 2019). Briefly, the LEQ is an informant (caregiver) based questionnaire that measures a range of educational, occupational, and social/leisure experiences during three life stages: young adulthood, mid-life, and late-life. Certain activities are considered to be undertaken primarily within one particular life phase, such as education (young), occupation (mid-life) and social engagement (late-life). Young adulthood questions uniquely focus on educational attainment (for example years of school completed and type of degree pursued), with post-secondary education being weighted higher. Mid-life questions obtain a complete occupational history alongside managerial experience and years worked at each position. Late-life questions predominantly revolve around social and intellectual activity undertaken during leisure time (for example how many social groups one belongs to and how often specific activities such as walking or strategy games are undertaken). All three life stage scores are then summed to produce a Total LEQ score.

### 2.3. Neuroimaging methods

All patients underwent a high-resolution research quality T1-weighted MRI scan. Scans were acquired using 3 very similar T1-weighted sequences as follows:

1. 3.0 Tesla SIEMENS TIM Trio scanner, 8 channel head coil, axial plane with repetition time = 1620ms, echo time = 3.87ms, slice thickness = 1.0mm, flip angle = 15 degrees, matrix = 192×256, in-plane resolution 0.98×0.98 mm.
2. 3.0 Tesla SIEMENS TIM Trio scanner, 64 channel head coil, sagittal plane with repetition time = 2300 ms, echo time = 2.95ms, slice thickness = 1.2mm, flip angle = 0 degrees, matrix = 256×240, in-plane resolution = 1.05×1.05 mm.
3. 3.0 Tesla SIEMENS Prisma scanner, 64 channel head coil, sagittal plane with repetition time 2400ms, echo time = 1.96ms, flip angle = 8 degrees, matrix = 320×320, slice thickness = 0.8mm, in-plane resolution = 0.8×0.8 mm.

Voxelwise cortical thickness maps were generated using Advanced Normalization Tools (ANTs), as previously reported (Tustison et al., 2014). Briefly, using antsCorticalThickness.sh, potential field bias was corrected using N4, brain extraction was performed using a template-based method, and six tissue class (cortex, deep gray matter, white matter, CSF, brainstem, and cerebellum) segmentation was performed using a combination of previously defined tissue priors as well as probabilistic tissue mapping (Tustison et al., 2010). Finally, easy_lausanne (https://github.com/mattcieslak/easy_lausanne) (Daducci et al., 2012) was run on our local template, which was created based upon data from the Open Access Series of Imaging Studies (Marcus et al., 2007) to create a standard cortical parcellation. The template parcellation was then spatially normalized to each participant’s native T1 space using the template-to-native T1 warps generated by ANTs, and then we calculated the mean CT in each region of interest (ROI) of the Lausanne125 scale, which we used for our analysis.

### 2.4. Statistical analyses

All analyses were conducted using R statistical software, utilizing packages: lmerTest (Kuznetsova et al., 2017) and PMCMR (Pohlert, 2014). All code is provided on an open-source platform (https://github.com/pennbindlab/SocialLEQ). Due to the large (n=219) number of ROIs, we set a significance threshold of family-wise error (FWE) corrected p < 0.05.

To identify regions of reduced cortical thickness (CT) relative to controls we performed a linear regression analysis within each ROI for the main effect of group controlling for T1-protocol (ROI Mean CT ∼ Group + T1Protocol) using baseline MRIs from the bvFTD cohort. ROIs identified to be significantly atrophied in the bvFTD cohort (n=121) compared to controls were included in a brain mask for subsequent analyses to limit our investigation of CR to regions affected by disease.

To evaluate associations between young, mid, and late-life LEQ and baseline CT, we performed linear regression analyses, covarying for age, sex, disease duration at MRI, and T1-protocol (ROI ∼ Late-Life LEQ Score + Age + Sex + Dx Duration + T1Protocol). All ROI ∼ LEQ models were conducted on the bvFTD group only. We also performed this analysis using the Total LEQ score as a main predictor of interest.

To evaluate whether LEQ scores relate to rate of longitudinal CT decline we performed linear mixed effects (LME) models. LMEs included fixed factors for age at baseline MRI, sex, T1-protocol, and random factor of disease duration at MRI date nested for each individual (ROI Mean CT ∼ (Dx Duration | id) + Sex + Age at Baseline MRI + T1Protocol). This model yielded an annualized fit for each individual that reflects the slope of CT decline within each ROI. We then performed linear regression analyses to relate the model generated CT slope to LEQ scores within each ROI. Similar to the baseline assessment, all longitudinal analyses were conducted on the bvFTD group only. We also performed this analysis using the Total LEQ score as a main predictor of interest.

### 2.5 Exploratory Social/Behavior Assessments

For social/behavioral measures, we used the Philadelphia Brief Assessment of Cognition (PBAC) (Libon et al., 2011) and the Social Behavior Observer Checklist (SBO) (Rankin, 2010) administered as part of the National Alzheimer’s Disease Coordinating Center’s Uniform Data Set (UDS) versions 2.0 and 3.0 (Weintraub et al., 2009) plus associated FTLD module versions 2.0 and 3.0 (Beekly et al., 2012). Briefly, trained testers observe participants over the course of a research visit and assign ratings on the PBAC and SBO based on social/behavioral comportment.

In addition to social/behavioral rating, the PBAC contains several neuropsychological tests evaluating domains of cognition (e.g. executive functioning - digit span backwards and letter-guided fluency). For cognitive assessments, we used composite subscores of the PBAC directed at two domains of cognition: the executive subscale, often affected in bvFTD, and the memory subscale as a control measure as memory is relatively preserved in bvFTD (Rascovsky et al., 2011)

## 3. RESULTS

### 3.1. Imaging results

There were widespread areas of cortical atrophy in frontal and temporal regions in bvFTD relative to healthy controls. (Figure 1A). At baseline, a higher late-life LEQ score was associated with reduced cortical atrophy in bilateral superior anterior temporal regions (Figure 1B). In our longitudinal analysis, a higher late-life LEQ score was associated with attenuated cortical loss in left insular cortex (Figure 1C). We did not observe baseline or longitudinal associations between any ROIs and the young-life, mid-life, or Total LEQ scores. A visual representation of effect size (Cohen’s F^2^, Selya et al., 2012) for all tested late-life LEQ regressions is shown in Supplementary Figure 1.

**Figure 1.**
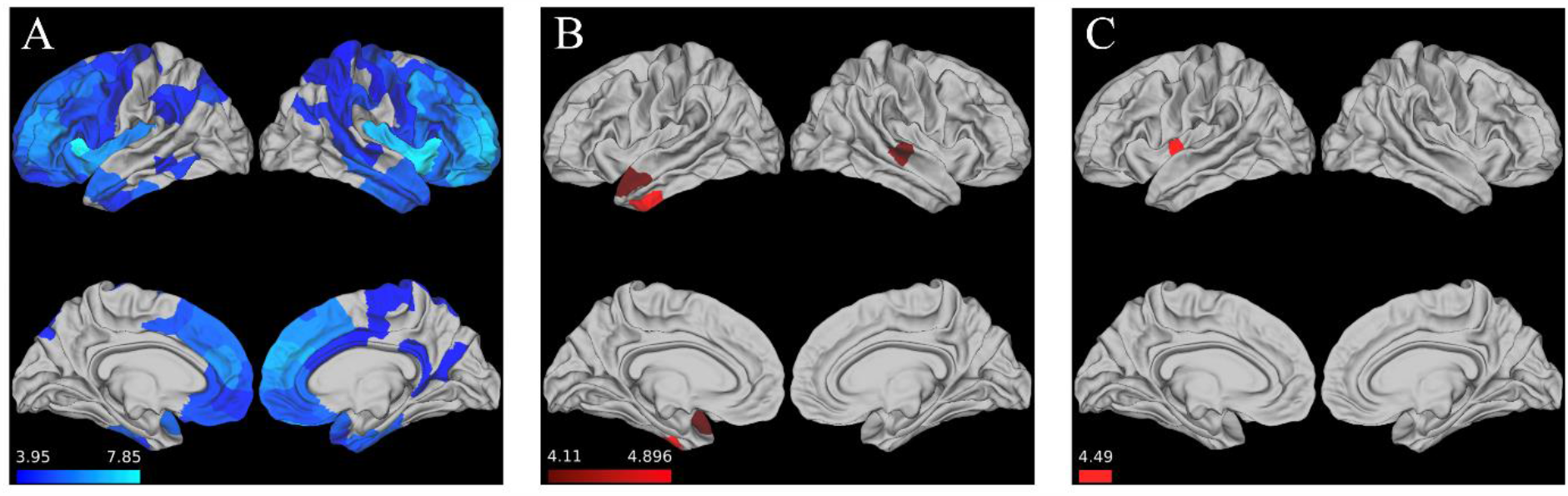
Late-Life LEQ relates to attenuated CT loss. A.) Areas with reduced baseline CT in bvFTD group vs. control group B.) Brain regions where at baseline late-life LEQ associated with reduced cortical atrophy C.) Brain regions where late-life LEQ associated with an attenuation of longitudinal cortical loss (scale bar for all images shows regression t-statistic)

### 3.2. Exploratory Cognitive Analysis

In an exploratory analysis, we sought to evaluate associations between late-life LEQ and social function, including the PBAC and SBO. In separate models, linear regression tested associations between PBAC and SBO and late-life LEQ score, covarying for age, sex, and disease duration at test (Assessment ∼ Late-Life LEQ + Age + Sex + Dx Duration). At baseline, a higher late-life LEQ score associated with less impairment on the PBAC behavior score (Figure 2: β=0.42, SE=0.08, p<0.05), but not on SBO. We did not observe significant associations between longitudinal PBAC and SBO measures and late-life LEQ (i.e. late-life LEQ did not moderate the rate of longitudinal decline). Lastly, we repeated these analyses using the PBAC executive and memory subscales and observed no significant associations at baseline or in longitudinal models. Descriptive statistics for all models tested are contained in Table 2.

**Table 2.**
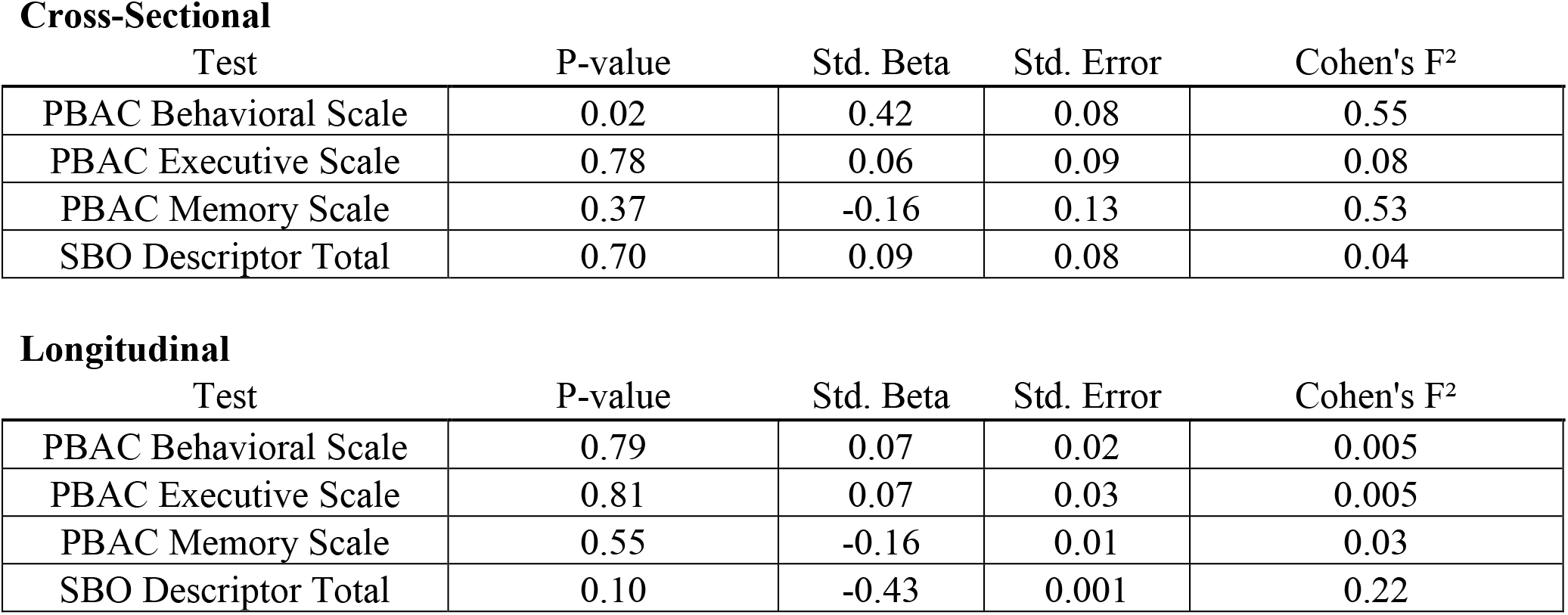
PBAC Subscale + SBO Descriptor associations with late-life LEQ

**Figure 2.**
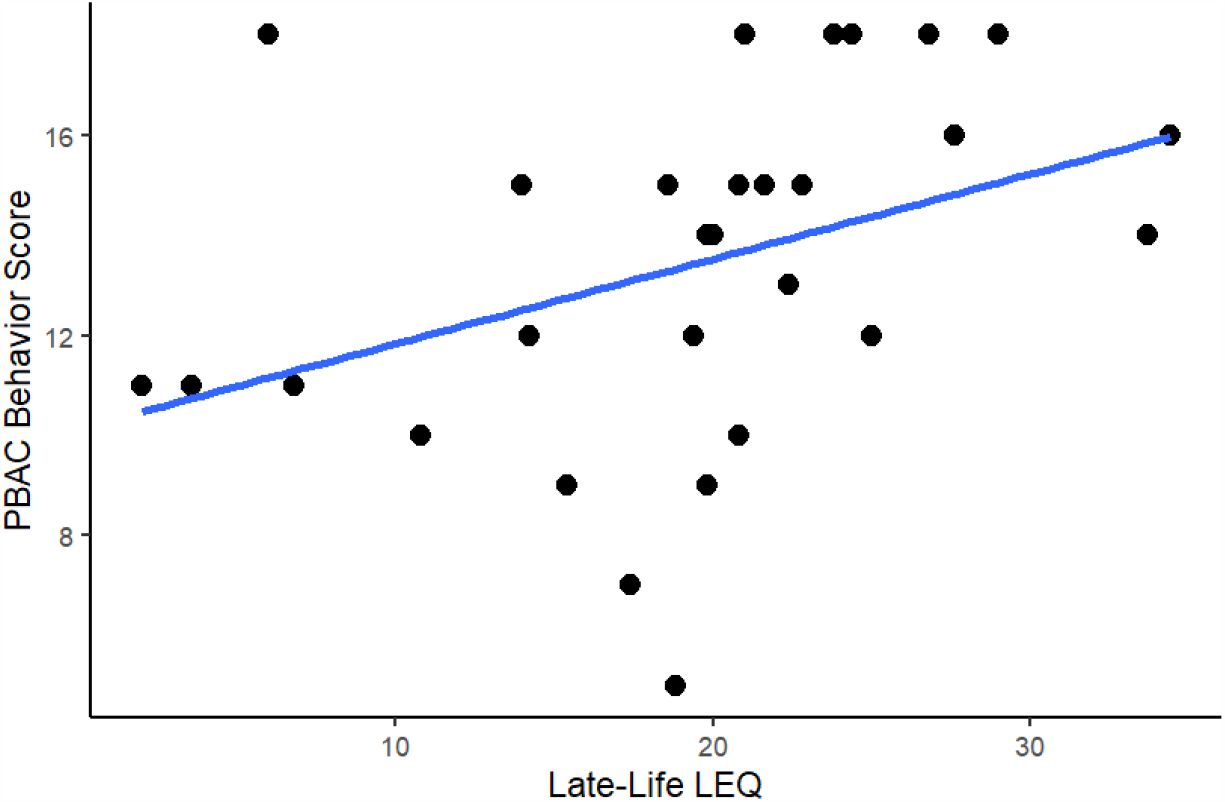
PBAC Behavioral Scale significantly relates to late-life LEQ (p<0.05) at baseline

## 4. DISCUSSION

In this study, we examined the effect of lifestyle factors such as education, occupational attainment, and social/leisure time activity across three life stages on brain structure in bvFTD. Our results suggest that social/leisure activity, reflected in late-life LEQ, is associated with relatively preserved gray matter in areas associated with social functioning, including superior anterior temporal regions and the insula. Consistent with CR theory, this work suggests that participation in such activities may contribute to a form of resilience in a common form of young-onset dementia that is marked by deficits in social comportment. However, it is important to consider the issue of reverse causality when interpreting our study. The LEQ is collected post-symptom onset, thus potential exists for our findings to reflect disease impairments rather than protective effects (i.e. does less degeneration in superior anterior temporal cortex lead to increased social engagement or does increased social engagement slow cortical degeneration in superior anterior temporal cortex). We attempted to account for this by controlling for disease severity in our analyses, however the possibility for this alternate interpretation exists. Our future work will be aimed at refining our measurement of social and leisure activity in order to clarify our results.

Our study is unique in the context of CR in bvFTD. Here we examine how social and leisure activity is associated with brain structure and social cognition. However, the majority of CR studies in bvFTD have been limited to education and occupation (Borroni et al., 2009; Dodich et al., 2018; Massimo et al., 2019, 2015; Placek et al., 2016; Spreng et al., 2011). In the present analysis, we also included young and mid-life LEQ scores, reflective of education level and occupational complexity, the most common proxies associated with CR. We did not observe a significant relationship between CT and these measures in our cross-sectional and longitudinal analyses. Previous studies in bvFTD have demonstrated that educational and occupational attributes are associated with integrity in frontal brain regions (Borroni et al., 2009; Dodich et al., 2018; Spreng et al., 2011). These studies suggest that individuals with high ‘reserve’ related to education and occupational attainment are better able to cope with neuronal degeneration in the frontal lobe, consistent with what is observed in traditional models of CR in AD. Null anatomic findings in our cohort may be related to differences in measurement of education and occupational attainment. For example, young and mid-life LEQ scores also consider other exposures such as hobbies and travel and thus may lack some specificity in measuring education and occupation. In a supplemental analysis, we evaluated our cross-sectional analyses using a voxelwise approach via the FMRIB Software Library’s randomise tool (Winkler et al., 2014). Similar to our ROI based method, we observed significant positive associations in the anterior temporal lobes for late-life LEQ (Supplementary Figure 2). Interestingly, we noted additional associations between Total LEQ and the right orbitofrontal cortex and the right dorsal anterior cingulate. This finding certainly warrants further investigation – it may be that the anatomical atlas we utilized (Lausanne125, Daducci et al., 2012) lacked sensitivity to detect some small scale regional relationships. The replication of our late-life LEQ associations is encouraging, however more work is needed to clarify our other null effects.

Despite null effects in our young and mid-life LEQ models, we cannot rule out the contribution of cognitive activities in young adulthood and mid-life to the observed association between late-life cognition and CT. Years of education has been shown to moderate the relationship between lifetime leisure activities and cognitive performance (Park et al., 2019), and it is feasible that in our cohort the resilience effect is not solely due to social and leisure activity. Indeed, the score of the life phases are reasonably collinear (Pearson R ranges from 0.16 – 0.44). In a supplemental analysis, we regressed late-life LEQ with early and mid-life and used the residuals as a representation of the independent effect of late-life cognition (i.e. the proportion of variance in the late-life LEQ unexplained by early and mid-life scores). Residuals (termed Partial late-life LEQ) were substituted for late-life LEQ in our models (Baseline: CT ∼ Residuals + Covariates, Longitudinal: Annualized Slope of CT Loss ∼ Residuals). Significant ROIs in this analysis had 100% overlap with our previous results – although results were less robust (p < 0.005, Supplementary Figure 3). As results did not survive a p-value correction, the possibility of Type I error exists, and thus we interpret these results as the independent effect of late-life cognition with caution.

Consistent with our hypothesis, brain regions found to be significantly associated with measures of social and leisure activity have been heavily implicated in social functioning. Imaging studies in healthy adults have shown that the insula as well as superior anterior temporal regions play vital roles in typical social interaction (Gogolla, 2017; Li et al., 2018). Indeed, the Social Context Network Model suggests that the integration of social contextual information from the external environment (e.g. body language and verbal prosody) necessary for social cognition is underpinned by a network of brain regions in the fronto-temporo-insular network, with direct overlap with our regions of interest (Ibañez and Manes, 2012). In bvFTD, difficulty processing external context clues such as facial expressions has been associated with disease in parahippocampal gyrus as well as temporal regions, amygdala, orbitofrontal cortex, and insula (De Winter et al., 2016; Kumfor et al., 2018; Woolley et al., 2015). Functional MRI studies have shown that bvFTD patients have reduced connectivity in the fronto-temporo-insular network, including our regions of interest, compared to controls (Agosta et al., 2013; Sedeño et al., 2016). Superior anterior temporal regions have also been highly implicated in high-level social functioning, with several studies investigating the intricacies of social cognition that rely on these brain regions (Rice et al., 2018; Skipper et al., 2011; Zahn et al., 2007). For example, work in bvFTD and semantic dementia patients has shown that superior anterior temporal regions play an important role in processing social concepts and this may be one mechanism for social and behavioral impairments in FTD (Pobric et al., 2015; Zahn et al., 2017). Our results provide some initial evidence that social engagement relates to cortical thickness in these areas, and more work is necessary to further specify how aspects of social activity and cognition relate to cortical degeneration in these highly complex regions.

Social engagement has been put forth as protective in aging and other neurodegenerative diseases such as AD. For example, two recent systematic reviews suggest that lower social engagement is associated with an increased risk of AD, and higher social engagement is protective in older adults (Hersi et al., 2017; Penninkilampi et al., 2018). Leisure time activities have also been found to improve clinical outcomes in genetic FTLD (Casaletto et al., 2020), and reduce incidence of other dementias such as AD (Scarmeas et al., 2001). While the mechanism by which such later life activities exert their effect on brain structure remains elusive, some have proposed that social stimulation might help in reducing the stress response which causes acceleration in neuronal degeneration (Hsiao et al., 2018). Resilience related factors may also include structural neuroanatomic factors that play a role in neural implementation of CR, such as supporting redundant or alternate brain networks for optimal performance. Of course we must also consider associated factors that are related to social and leisure activities which may include greater physical and cognitive activity, and more in-depth research in this area is necessary to disentangle potential interactions among these factors and to elucidate how they attenuate decline. While our exploratory behavioral analysis provided some support for a beneficial effect of social and leisure activity on social behavior at the baseline assessment, we did not observe a similar association on longitudinal decline. Our results should be interpreted keeping in mind that the social/behavioral measures used in this study may be subject to confounds. For example, ratings are generated based upon patient behavior during a research session, during which one’s behavior is likely different than in their daily environment. Furthermore, longitudinal scales were completed by several different raters, potentially introducing more potential for measurement error. With this in mind, our behavioral analysis provides some evidence that the effect of social and leisure activity on CT is specific to ameliorating social and behavioral impairments; however, more work needs to be done using additional behavioral measures to investigate this effect on behavior in bvFTD.

Our results provide new information about CR in bvFTD. Since current treatments for bvFTD are aimed at symptom management (Logroscino et al., 2019), our results provide potential interventions such as increasing social stimulation and participating in leisure time activity for mitigation of social deficits. However, our study should be interpreted with some caveats. First, our longitudinal sample was limited by the availability of follow up data, and with a larger sample this effect could be evaluated more thoroughly. For example, follow-up time and number of datapoints collected can vary due to reasons such as illness, death, unwillingness to participate due to disease progression, and transportation issues. Therefore, to ensure each subject was represented in longitudinal models equally, we chose to perform linear mixed effects modeling to extract individualized slopes of decline and relate this metric to late-life LEQ, similar to previous work (Placek et al., 2020). However, such a method has the potential to be biased by shrinkage towards the mean. In a post-hoc analysis, we pursued an alternate approach to modeling the effect of late-life LEQ on longitudinal decline – modeling the effect of the interaction between LEQ score and time elapsed from baseline on CT (CT ∼ LEQ*TimeElapsed + Age at Baseline MRI + Sex + Dx Duration + T1Protocol). We observed an attenuation of cortical loss in the right anterior temporal lobe and insular cortex (Supplementary Figure 4).

A second limitation of our study was the lack of available PBAC data within our neuroimaging cohort. A “gold standard” approach to testing resilience effects is to test the interaction between lifetime experience and CT on cognition to question whether increased lifetime experience can moderate the brain-behavior relationship. This is well reflected in (Casaletto et al., 2020), however to test such an interaction in our patient cohort requires both MRI and PBAC to be available in all cases, which unfortunately was not available to us. Future work is necessary to address such an interaction effect, and in a prospective study we aim to investigate this further.

Finally, as mentioned above, the LEQ is collected post-symptom onset, and is a more subjective measure than other CR proxies such as education and occupation, thus potential exists for our findings to reflect disease impairments rather than protective effects. Our future work will be aimed at refining our measurement tools to validate that our findings indeed reflect the protective effects of social and leisure activity. Furthermore, as bvFTD is only a single phenotype of FTLD spectrum disorders, we hope to expand our study to other FTLD subtypes as well as individuals with multiple concurrent syndromes in order to further investigate how CR affects progression in this clinically and pathologically heterogeneous disorder.

## 5. CONCLUSIONS

With these caveats in mind, this study suggests that increased participation in social and leisure activities may contribute to a form of resilience to bvFTD in areas associated with social cognition, adding to the burgeoning CR literature in non-Alzheimer’s dementias. With the majority of previous studies on CR in bvFTD focusing on education and occupation, we provide a novel finding of activities that have the potential to mitigate disease progression.

## Data Availability

Anonymized data will be shared by a reasonable request from any qualified investigator.

https://github.com/pennbindlab/SocialLEQ

## Abbreviations

(CR): Cognitive reserve
(bvFTD): Behavioral Variant Frontotemporal Degeneration
(LEQ): Lifetime of Experiences Questionnaire
(CT): Cortical Thickness
(MRI): Magnetic Resonance Imaging
(AD): Alzheimer’s Disease
(FTLD): Frontotemporal Lobar Degeneration
(FTDC): Penn Frontotemporal Degeneration Center
(ROI): region of interest
(PBAC): Philadelphia Brief Assessment of Cognition
(SBO): Social Behavior Observer Checklist
(UDS): National Alzheimer’s Disease Center Uniform Data Set
(ANTs): Advanced Normalization Tools

## ACKNOWLEDGEMENTS

The authors thank the research study participants, without whom this would not be possible.

## Author contributions

**NGK**, Conceptualization, Methodology, Software, Formal analysis, Validation, Writing – Original Draft, Project Administration. **JB**, Conceptualization, Software, Formal analysis, Writing – Original Draft. **DGW**, Supervision. **JSP**, Supervision, Writing – Review & Editing. **KAC**, Data Curation, Writing – Review & Editing. **CAO**, Visualization, Writing – Review & Editing. **CTM**, Supervision, Conceptualization, Methodology, Writing – Review & Editing. **LM**, Supervision, Funding acquisition, Writing – Review & Editing, Conceptualization, Methodology

## Patient consent for publication

All participants and legally responsible next of kin completed written informed consents for the MRI protocols and neuropsychological testing, which are approved by University of Pennsylvania’s Institutional Review Board.

## Data availability statement

Anonymized data will be shared by a reasonable request from any qualified investigator.

## APPENDIX A. SUPPLEMENTARY FIGURES

**Supplementary Figure 1.**
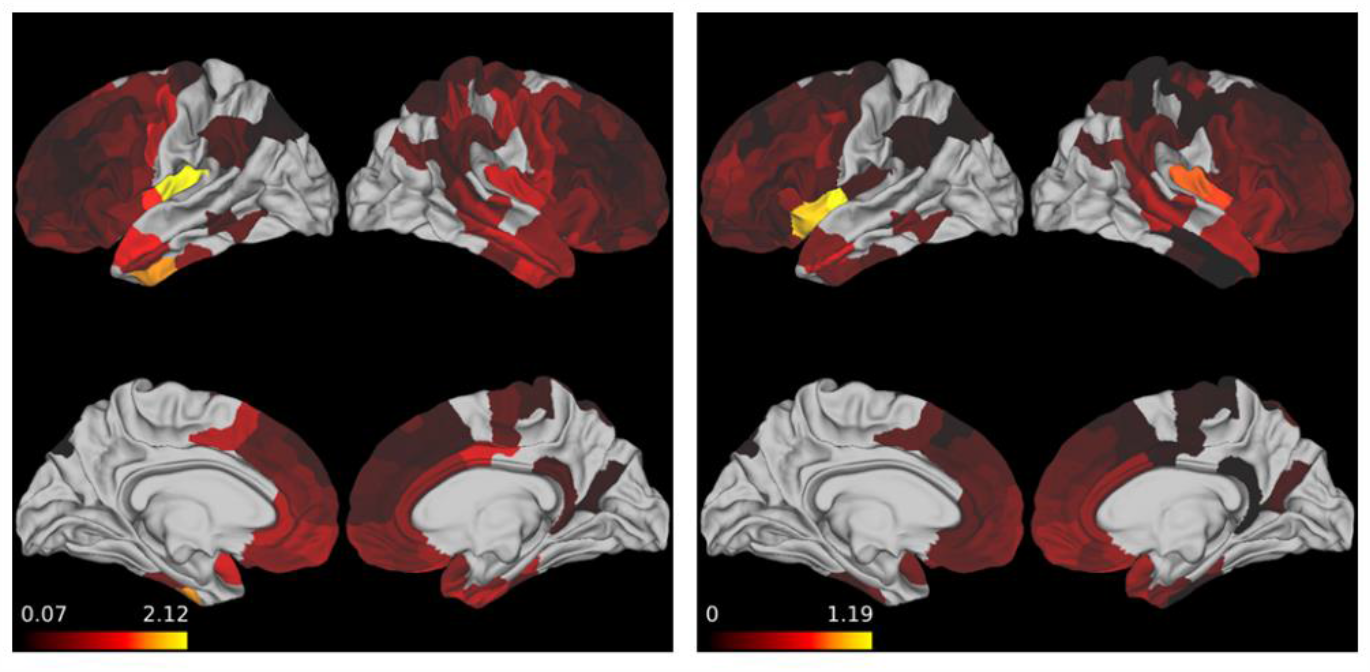
Heatmap of Cohen’s F^2^ for tested multiple linear regressions <u>Note:</u> Left panel – Cross-sectional analysis, Right panel – Longitudinal analysis

**Supplementary Figure 2.**
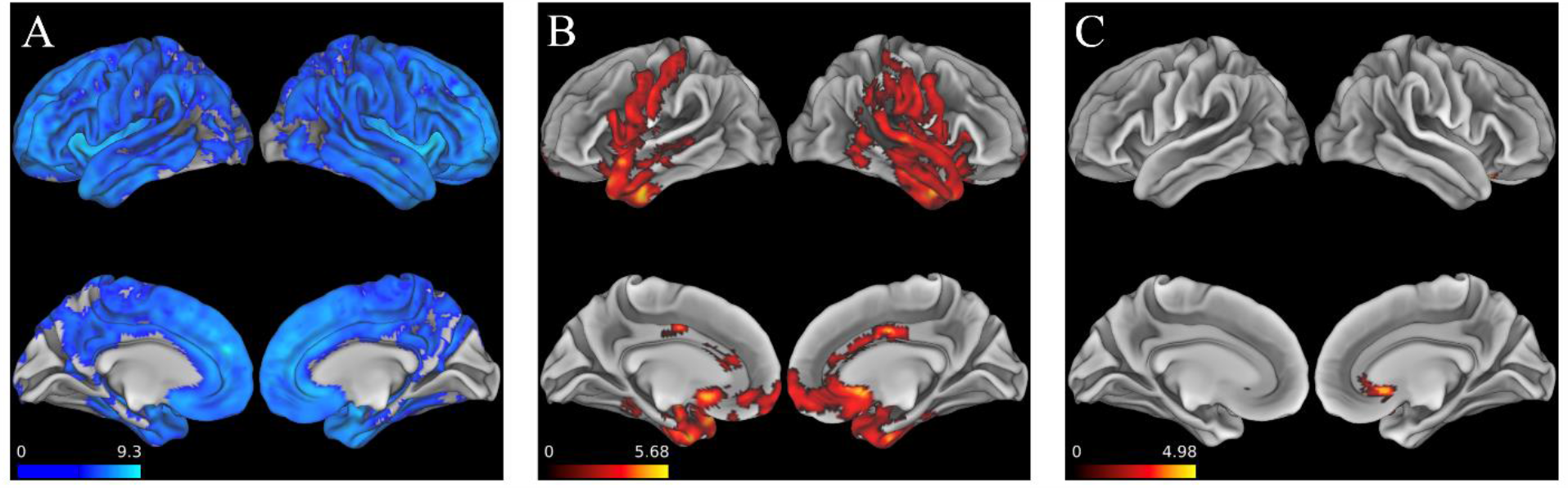
Voxelwise analysis A.) Areas of cortical atrophy in bvFTD cohort vs. control group B.) Areas where late-life LEQ significantly related to reduced cortical atrophy at baseline C.) Areas where Total LEQ significantly related to reduced cortical atrophy. All analyses were performed with a significance threshold of FWE corrected p < 0.05 and a cluster size threshold of 50 voxels.

**Supplementary Figure 3.**
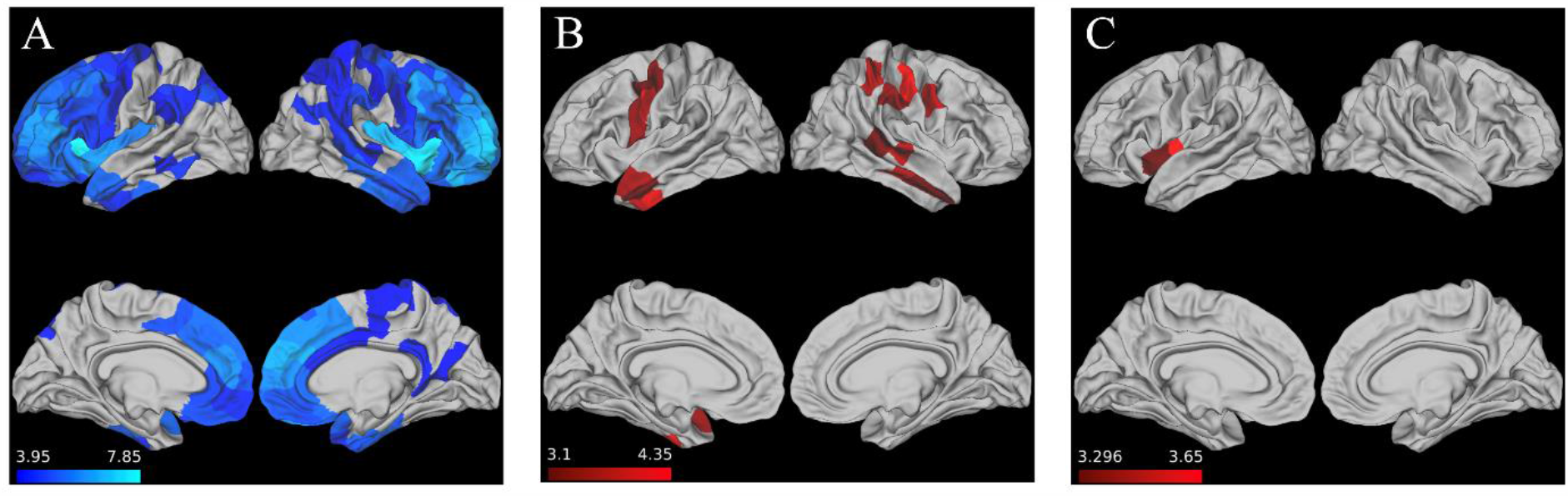
Partial Late-life LEQ derived from residualized model associations with CT. A.) Areas of significant (FWE p < 0.05) cortical atrophy in bvFTD cohort vs. control group (Note – same as reported in **Figure 1**) B.) Areas where partial late-life LEQ significantly (p < 0.005) related with reduced cortical atrophy at baseline C.) Areas where partial late-life LEQ significantly (p < 0.005) associated with an attenuation of cortical loss.

**Supplementary Figure 4.**
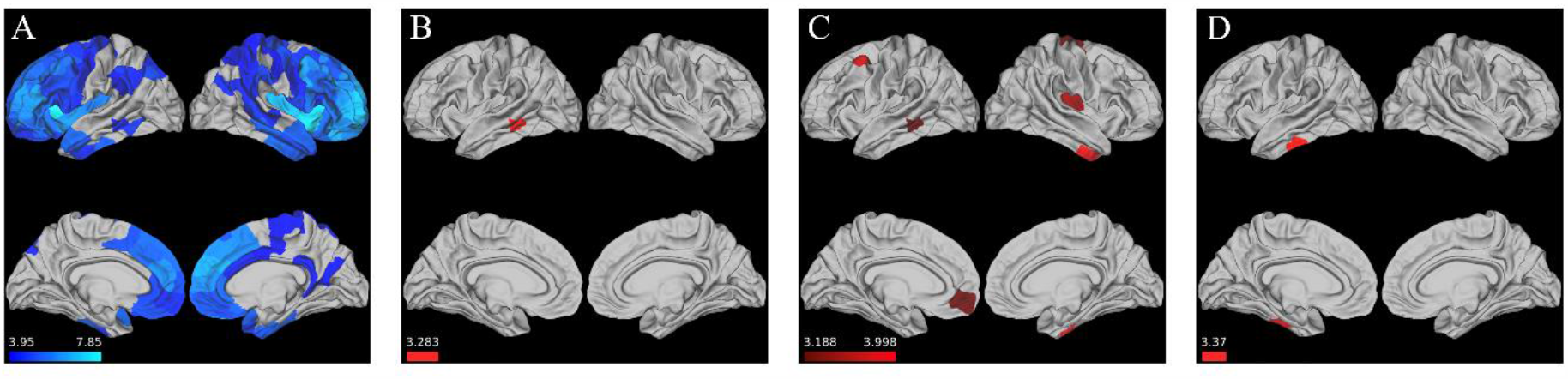
LEQ * Time Interaction shows attenuation of cortical loss A.) Areas of significant (FWE corrected p < 0.05) cortical atrophy in bvFTD cohort vs. control group (Note – same as reported in **Figure 1**) B.) Area where mid-life LEQ significantly (p < 0.005) associated with an attenuation of cortical loss C.) Area where late-life LEQ significantly (p < 0.005) associated with an attenuation of cortical loss D.) Area where Total LEQ significantly (p < 0.005) associated with an attenuation of cortical loss.

